# No Clear Benefit to the Use of Corticosteroid as Treatment in Adult Patients with Coronavirus Disease 2019 : A Retrospective Cohort Study

**DOI:** 10.1101/2020.04.21.20066258

**Authors:** Dan Wang, Juan Wang, Qunqun Jiang, Juan Yang, Jun Li, Chang Gao, Haiwei Jiang, Lintong Ge, Yongming Liu

**Affiliations:** Department of Neurology, The Third People’s Hospital of Hubei Province, Jianghan University, Wuhan, China; Department of Radiology, Tongji Hospital, Huazhong University of Science and Technology, Wuhan, China; Department of Infectious Diseases, Zhongnan Hospital of Wuhan University, Wuhan, China

**Keywords:** COVID-19, SARS-CoV-2, Corticosteroid, Cohort study, Outcome

## Abstract

**Background:** Coronavirus disease 2019 (COVID-19) is becoming an increasing global health issue which has spread across the globe. We aimed to study the effect of corticosteroids in the treatment of adult inpatients with COVID-19.

**Methods:** A retrospective cohort of 115 consecutive adult COVID-19 patients admitted to The Third People’s Hospital of Hubei Province between Jan 18, 2020, and Feb 28, 2020 was analysed to study the effectiveness of corticosteroid. They were categorized according to whether or not corticosteroid therapy was given, and compared in terms of demographic characteristics, clinical features, laboratory indicators and clinical outcomes. The primary endpoint was defined as either mortality or intensive care unit (ICU) admission. Known adverse prognostic factors were used as covariates in multiple logistic regressions to adjust for their confounding effects on outcomes.

**Results:** Among 115 patients, 73 patients (63.5%) received corticosteroid. The levels of age, C-reactive protein, D-dimer and albumin were similar in both groups. The corticosteroid group had more adverse outcomes (32.9% vs. 11.9%) and statistically significant differences were observed (p=0.013). In multivariate analysis, corticosteroid treatment was associated with a 2.155-fold increase in risk of either mortality or ICU admission, although not statistically significant.

**Conclusion:** No evidence suggests that adult patients with COVID-19 will benefit from corticosteroids, and they might be more likely to be harmed with such treatment.

## Introduction

The epidemic of coronavirus disease 2019 (COVID-19), an emerging infectious respiratory disease caused by severe acute respiratory syndrome coronavirus 2 (SARS-CoV-2)[1], started in Wuhan, China in november 2019 and swept over multiple countries and regions worldwide. With regards to the treatment of COVID-19, no specific approaches so far are available[2].

The use of corticosteroids remains controversial. During the outbreaks of severe acute respiratory syndrome (SARS)-CoV and middle east respiratory syndrome (MERS)-CoV, corticosteroids were widely used[3]. Similarly, physicians tend to use corticosteroids in critically ill patients with COVID-19 in addition to other therapeutics[4]. However, someone had been sceptical about its curative effect, especially in higher dosages, which carry long-term side effects. According to current World Health Organization (WHO) guidance[5], Russell[2] recommend that corticosteroids should not be used in SARS-CoV-2-induced lung injury or shock unless indicated for another reason.

We performed a retrospective cohort study on the effect of corticosteroid in a group of patients (age≥18years) with COVID-19, aimed to provide a scientific and rational medical decision-making basis for clinicians.

## Methods

### Study design and participants

This single-center, retrospective cohort study included individuals who were diagnosed with laboratory-confirmed COVID-19 consecutively between Jan 18, 2020, and Feb 28, 2020, at The Third People’s Hospital of Hubei Province. Eligible patients were those aged 18 years or older infected with SARS-CoV-2, which was diagnosed by real-time reverse-transcriptase polymerase-chain-reaction (RT-PCR) assay for nasal or pharyngeal swab specimens[6].

All patients with COVID-19 enrolled in this study were diagnosed and classified according to the new coronavirus pneumonia diagnosis and treatment plan (trial version 7) developed by the National Health Committee of the People’s Republic of China[7]. In this study, patients with mild or moderate symptoms were classified as noncritical, and patients with severe or critical symptoms were classified as critical.

### Procedures

All patients were treated in the SARS-CoV-2 infection isolation wards. Their oxygen saturation and vital signs were closely monitored. Chest CT scans, laboratory indicators including serial haematological, biochemical tests, coagulation parameters and inflammation-related factors were performed.

All patients were empirically treated with intravenous moxifloxacin at 0.4g per day for atypical and typical pneumonia, arbidol was given at a dose of 0.2g every 8 h, together with intravenous ribavirin at 0.5g every 12h as antiviral therapy. Corticosteroid therapy was commenced at the discretion of the attending clinicians after informed consent had been obtained from the patients themselves or their next of kin. The options for the corticosteroid regimen were: pulse intravenous methylprednisolone 0.5-1.0g per day for 2-3 days; or intravenous methylprednisolone at 1-3 mg/kg per day for 3-10 days. Other treatment options included immunoglobulin, interferon-alpha, traditional chinese medicine or any combination of the above.

### Outcome

The primary endpoint was the occurrence of an adverse outcome, which was defined as either mortality or intensive care unit (ICU) admission.

### Data Collection

115 consecutive adult patients fulfilling the criteria of COVID-19 were studied. Their demographic characteristics, epidemiological history, comorbidity, clinical features, chest imaging results, laboratory indicators, corticosteroid treatment options and clinical outcomes were obtained from electronic medical records. The data collection forms were reviewed independently by two experienced physicians.

### Statistical analysis

Categorical variables were given as frequency rates and percentages; continuous variables were defined using mean, median, and interquartile range (IQR) values. The Kolmogorov-Smirnov test was used to verify the normality of distribution of continuous variables. The independent sample t test or the Mann-Whitney U test was used for the continuous variables and the Chi-Square test or Fisher-Exact test for categorical variables. The known adverse prognostic factors for COVID-19 mortality including old age[8-10], male[10-12], presence of comorbidities[10], high D-dimer levels[8, 13], high lactate dehydrogenase levels[13, 14], high C-reactive protein levels[9], low lymphocyte count levels[14-17], low serum potassium[18] and albumin[14] were used as covariates in multiple logistic regressions to adjust for their confounding effects on adverse outcomes. Statistical analyses were performed using SPSS 24.0 (SPSS Inc, Chicago, IL, USA). A 2-tailed P<0.05 was considered as statistically significant.

## Results

A total of 136 patients older than 18 years old were diagnosed with COVID-19 infection between Jan 18, 2020, and Feb 28, 2020. No abnormal chest imaging was present in 8 patients. Thirteen patients were transferred to other hospitals. After excluding ineligible patients, 115 patients were included in the study **(Figure 1)**.

**Figure 1.**
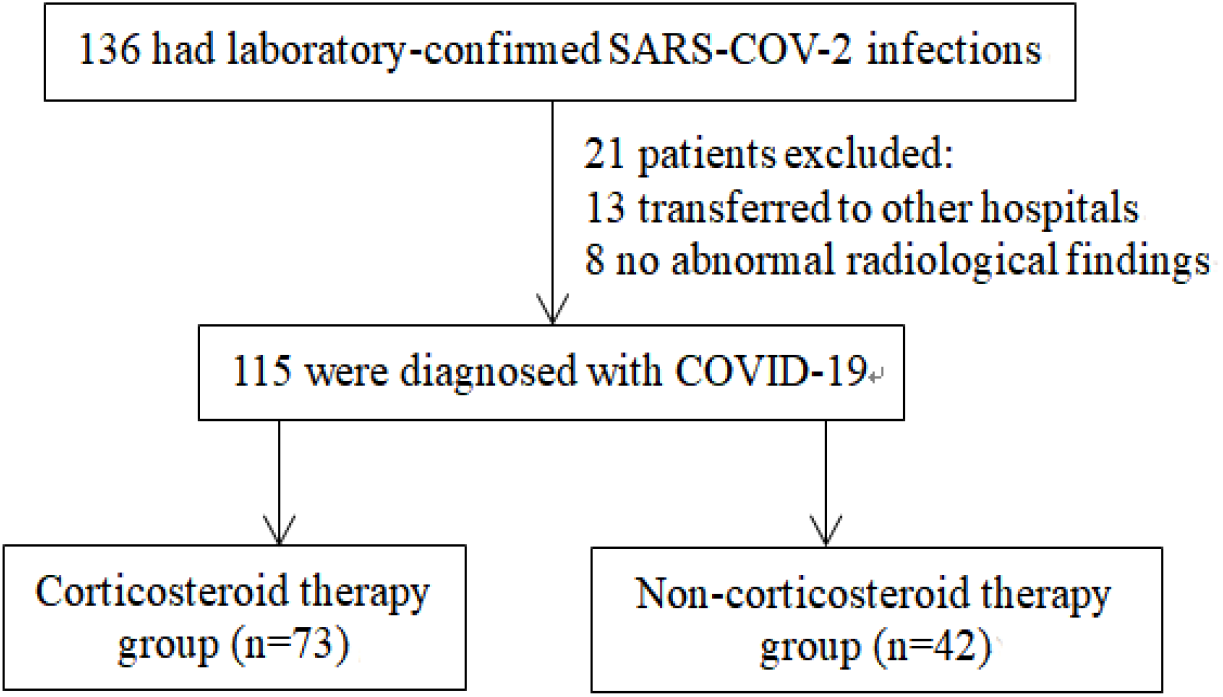
Study profile.

There were 60 noncritical and 55 critical cases among 115 enrolled COVID-19 patients. The median age of all patients was 59 years (IQR, 40-67), and 58 (50.4%) were men. Of the 115 patients, 42 (36.5%) had 1 or more comorbidities. The most common symptoms at initial stage of illness were fever (111[96.5%]), fatigue (72 [62.6%]) and dry cough (63 [54.8%]). Less common symptoms were nausea or vomiting and headache. Compared with noncritical cases, critical cases were significantly older (median age, 65 years [IQR, 56-69] vs 46 years [IQR, 33-62] ; P <0.001) and were more likely to have underlying comorbidities. Seventy-three cases (63.5%) received corticosteroid of various dosages. Steroid therapy for patients was heterogeneous, with 31 (51.7%) noncritical patients and 42 (76.4%) critical patients receiving corticosteroids (**Table 1**).

**Table 1.** Demographics and baseline characteristics of patients with COVID-19

|  | No. (%) |  |  | *p* value <sup>a</sup> |
| --- | --- | --- | --- | --- |
|  | Total (n=115) | Noncritical(n=60) | Critical (n=55) |  |
| Age, median (IQR), y | 59 (40-67) | 46 (33-62) | 65 (56-69) | 0.000 |
| Male | 58 (50.4%) | 24 (40.0%) | 34 (61.8%) | 0.019 |
| Comorbidities | 42 (36.5%) | 9 (15.0%) | 33 (60.0%) | 0.000 |
| Hypertension | 30 (26.1%) | 4 (6.7%) | 26 (47.3%) | 0.000 |
| Cardiovascular disease | 14 (12.2) | 4 (6.7%) | 10 (18.2%) | 0.059 |
| Diabetes | 12 (10.4%) | 4 (6.7%) | 8 (12.3%) | 0.167 |
| Signs and symptoms |  |  |  |  |
| Fever | 111 (96.5%) | 59 (98.3%) | 52 (94.5%) | 0.550 |
| Fatigue | 72 (62.6%) | 34 (56.7%) | 38 (69.1%) | 0.169 |
| Dry cough | 63 (54.8%) | 30 (50.0%) | 33 (60%) | 0.282 |
| Anorexia | 49 (42.6%) | 18 (30.0%) | 31 (56.4%) | 0.004 |
| Chest tightness | 54 (47.0%) | 26 (43.3%) | 28 (50.9%) | 0.416 |
| Myalgia | 42 (36.5%) | 16 (26.7%) | 26 (47.3%) | 0.022 |
| Chill | 29 (25.2%) | 17 (28.3%) | 12 (21.8%) | 0.422 |
| Dyspnea | 28 (24.3%) | 10 (16.7%) | 18 (32.7%) | 0.045 |
| Pharyngalgia | 23 (20%) | 11 (18.3%) | 12 (21.8%) | 0.641 |
| Nausea or Vomiting | 13 (11.3%) | 7 (11.7%) | 6 (10.9%) | 0.898 |
| Headache | 7 (6.1%) | 3 (5.0%) | 4 (7.3%) | 0.905 |
| White blood cell count, median (IQR), $\times 10^9/L$ | 3.3 (3.2-5.7) | 3.6 (3.2-4.1) | 4.5 (3.1-7.2) | 0.016 |
| Lymphocyte count, median (IQR), $\times 10^9/L$ | 0.9 (0.6-1.2) | 1.0 (0.7-1.3) | 0.7 (0.6-1.0) | 0.002 |
| Serum potassium, | 3.5 (3.2-3.6) | 3.5 (3.3-3.6) | 3.4 (3.2-3.6) | 0.063 |
| median (IQR), mmol/L |  |  |  |  |
| Creatinine, | 61 (50-74) | 59 (51-71) | 67 (50-79) | 0.225 |
| median (IQR), μmol/L | | | | |
| Lactate dehydrogenase, | 255 (168-354) | 219 (155-271) | 342 (249-476) | 0.000 |
| median (IQR), U/L |  |  |  |  |
| Serum amyloid A, | 209 (30.2-654.4) | 39.4 (12.9-139.3) | 582.3 (260.7-1074.7) | 0.000 |
| median (IQR), mg/L |  |  |  |  |
| C-reactive protein, | 21.7 (5.0-71.2) | 9.7 (4.7-33.2) | 64.7 (13.9-101.9) | 0.000 |
| median (IQR), mg/L |  |  |  |  |
| ESR, median (IQR), | 34.1 (18.0-61.1) | 25.8 (16.7-48.4) | 43.0 (21.6-75.3) | 0.009 |
| mm/h |  |  |  |  |
| CK-MB, median (IQR), | 12.3 (9.5-18.2) | 12.2 (9.6-16.2) | 12.3 (9.5-20.3) | 0.404 |
| U/L |  |  |  |  |
| Albumin, median (IQR), | 36.7 (32.0-39.6) | 39.0 (37.0-40.7) | 32.0 (30.2-34.3) | 0.000 |
| g/L |  |  |  |  |
| D-dimer, median (IQR), | 0.5 (0.3-1.8) | 0.4 (0.2-0.5) | 1.3 (0.4-2.8) | 0.000 |
| ug/mL |  |  |  |  |
| Receive corticosteroid | 73 (63.5%) | 31 (51.7%) | 42 (76.4%) | 0.003 |
| therapy |  |  |  |  |

Abbreviations: IQR, interquartile range; COVID-19, coronavirus disease 2019; ESR, erythrocyte sedimentation rate; CK-MB, creatine kinase-muscle and brain type; <sup>a</sup> *P* values indicate differences between noncritical and critical. *P*<0.05 was considered statistically significant.

Twenty-nine (25.2%) of our patients reached the adverse outcomes of ICU admission or mortality. Patients with adverse outcomes were older and had more comorbidities. Their lymphocyte counts and albumin levels were lower, their D-dimer, C-reactive protein, and lactate dehydrogenase levels were higher (**Table 2**).

**Table 2.** Adverse prognostic factors associated with either ICU admission or mortality

|  | Median (IQR) |  | *p* value<sup>a</sup> |
| --- | --- | --- | --- |
|  | ICU admission or mortality (n=29) | Uncomplicated (n=86) |  |
| Age, y | 68 (61-71) | 51 (35-65) | 0.000 |
| Male, No. (%) | 16 (55.2%) | 42 (48.8%) | 0.555 |
| Comorbidities, No. (%) | 19 (65.5%) | 26 (30.2%) | 0.001 |
| Lymphocyte count, $\times 10^9$/L | 0.7 (0.5-0.9) | 0.9 (0.7-1.2) | 0.001 |
| Serum potassium, mmol/L | 3.3 (3.2-3.5) | 3.5 (3.3-3.7) | 0.084 |
| Albumin, g/L | 32 (30-35) | 38 (33-40) | 0.000 |
| D-dimer, ug/mL | 2.0 (0.8-10.3) | 0.4 (0.3-0.9) | 0.000 |
| Lactate dehydrogenase, U/L | 398 (272-521) | 244 (162-296) | 0.000 |
| C-reactive protein, mg/L | 84.1 (49.8-119.0) | 13.2 (4.8-45.4) | 0.000 |

Abbreviations: ICU: intensive care unit; IQR, interquartile range; <sup>a</sup> *P* values indicate differences between ICU admission or mortality and uncomplicated. *P*<0.05 was considered statistically significant.

When patients were categorized according to whether corticosteroid treatment was given, and compared in terms of outcomes, as well as age comorbidities and other established adverse prognostic factors[8-10, 12-16, 18], the corticosteroid group was found to had more comorbidities, lower lymphocyte count and higher LDH. The age, sex, serum potassium, albumin, D-dimer and C-reactive protein level were similar in both groups. The corticosteroid group had more adverse outcomes (32.9% vs. 11.9%), and statistically significant differences were observed (p=0.013) (**Table 3**).

**Table 3.**
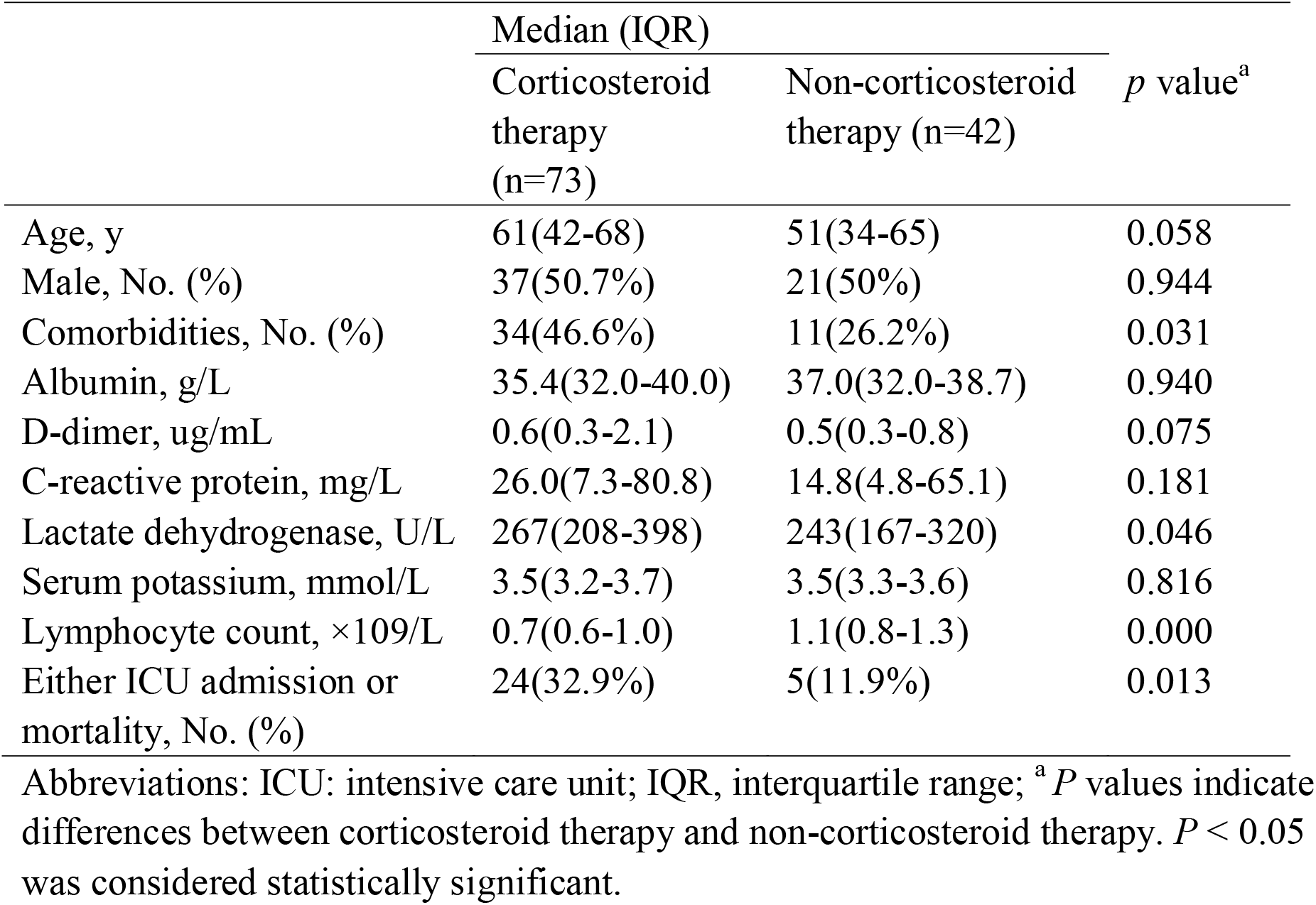
Comparison of characteristics, adverse prognostic factors and outcomes between corticosteroid therapy group and non-corticosteroid therapy group

|  | Median (IQR) |  | *p* value<sup>a</sup> |
| --- | --- | --- | --- |
|  | Corticosteroid therapy (n=73) | Non-corticosteroid therapy (n=42) |  |
| Age, y | 61(42-68) | 51(34-65) | 0.058 |
| Male, No. (%) | 37(50.7%) | 21(50%) | 0.944 |
| Comorbidities, No. (%) | 34(46.6%) | 11(26.2%) | 0.031 |
| Albumin, g/L | 35.4(32.0-40.0) | 37.0(32.0-38.7) | 0.940 |
| D-dimer, ug/mL | 0.6(0.3-2.1) | 0.5(0.3-0.8) | 0.075 |
| C-reactive protein, mg/L | 26.0(7.3-80.8) | 14.8(4.8-65.1) | 0.181 |
| Lactate dehydrogenase, U/L | 267(208-398) | 243(167-320) | 0.046 |
| Serum potassium, mmol/L | 3.5(3.2-3.7) | 3.5(3.3-3.6) | 0.816 |
| Lymphocyte count, ×10<sup>9</sup>/L | 0.7(0.6-1.0) | 1.1(0.8-1.3) | 0.000 |
| Either ICU admission or mortality, No. (%) | 24(32.9%) | 5(11.9%) | 0.013 |

Abbreviations: ICU: intensive care unit; IQR, interquartile range; <sup>a</sup>*P* values indicate differences between corticosteroid therapy and non-corticosteroid therapy. *P* < 0.05 was considered statistically significant.

Multivariate analysis was performed to adjust for the confounding effect of disease severity. Patients who were treated with corticosteroid were found to have a 2.155-fold increased risk of either ICU admission or mortality, although not statistically significant (**Table 4**).

**Table 4.** Adverse prognostic factors associated with either ICU admission or mortality by multivariate analysis

|  | Adjusted odds ratio | 95% CI | *p* value |
| --- | --- | --- | --- |
| Age | 1.061 | 1.002-1.124 | 0.043 |
| C-reactive protein | 1.022 | 1.007-1.037 | 0.005 |
| Corticosteroid therapy | 2.155 | 0.493-9.427 | 0.308 |

## Discussion

Acute respiratory distress syndrome (ARDS) and acute lung injury are partly induced by host immune responses[4]. Corticosteroids could suppress inflammation, but the pathogen clearance and immune response are also inhibited at the same time[19]. Systemic inflammation has been associated with adverse outcomes in SARS-CoV infection[20] and inflammation also persists after viral clearance[21]. In theory, corticosteroid administration can be applied to suppress lung inflammation.

However, in a meta-analysis[3] of corticosteroid use in SARS patients, only four studies provided conclusive data and indicated harm including psychosis[22], viraemia[19], diabetes and avascular necrosis[23, 24]. In a retrospective study reporting on 309 adults who were critically ill with MERS, patients given corticosteroids were more likely to obtain vasopressors, mechanical ventilation and renal replacement therapy[25]. A 2019 systematic review and meta-analysis[26] in influenza revealed increased mortality in patients who were given corticosteroids.

This report, to our knowledge, is the first cohort study to describe a role for corticosteroid in patients with COVID-19. In our series, twenty-nine (25.2%) patients reached the adverse outcomes of ICU admission or mortality. The adverse prognostic indicators were similar to those in other studies, including old age[8-10] and high C-reactive protein levels[9]. Despite similar age and C-reactive protein levels, the corticosteroid group had more adverse outcomes (32.9% vs. 11.9%) and statistically significant differences were observed (p=0.013). In multivariate analysis, corticosteroid treatment was associated with a 2.155-fold increase in risk of adverse outcome, although not statistically significant. This may be due to a small sample size and multicollinearity, or the characteristics of the sample. Whatever the reason, no clinical data exists to indicate that net benefit is derived from corticosteroids in the treatment of COVID-19.

This study has several limitations. First, the sample size is relatively small. The results could have been biased by some unknown confounding factors. Second, we did not stratify the outcomes according to various corticosteroid dosages, methods and lasting periods.

## conclusion

No evidence suggests that adult patients with COVID-19 will benefit from corticosteroids in this study. Considering the severe complications triggered by glucocorticosteroid such as psychosis[22], viraemia[19], diabetes and avascular necrosis[23, 24], the patients might be more likely to be harmed with such treatment. More studies are need to evaluate the clinical curative effect, as well as the appropriate dosages and duration of corticosteroid treatment in COVID-19.

## Data Availability

The datasets used and/or analyzed during the current study are available from the corresponding author on reasonable request.

## Funding

None.

## Conflict of interest

The authors have declared that no competing interest exists.

## Author Contributions

Dan Wang was responsible for the conception and design of the study. Dan Wang, Qunqun Jiang and Juan Wang were responsible for acquisition and analysis of data; furthermore, Dan Wang, Jun Li and Chang Gao were in charge of statistical analysis. Dan Wang and Juan Wang took part in drafting the manuscript; Lintong Ge, Haiwei Jiang and Yongming Liu revised and approved the final version of the manuscript. All authors read and approved the final manuscript.

## Ethics approval and consent to participate

This study was approved by the ethical committee of The Third People’s Hospital of Hubei Province. Because of the infectivity and the exploration urgency for COVID-19, written informed consent was waived and oral consent was obtained.

## Abbreviations

COVID-19: coronavirus disease 2019
SARS-CoV-2: severe acute respiratory syndrome coronavirus 2
WHO: World Health Organization
ARDS: acute respiratory distress syndrome
ICU: intensive care unit
RT-PCR: real-time reverse-transcriptase polymerase-chain-reaction
MERS: middle east respiratory syndrome
IQR: interquartile range
CK-MB: Creatine kinase-muscle and brain type
ESR: Erythrocyte sedimentation rate.

## Notes

### Competing Interest Statement

The authors have declared no competing interest.

